# Estimates of outbreak-specific SARS-CoV-2 epidemiological parameters from genomic data

**DOI:** 10.1101/2020.09.12.20193284

**Authors:** Timothy G. Vaughan, Jérémie Sciré, Sarah A. Nadeau, Tanja Stadler

**Author notes:** TGV, JS, SAN and TS designed research; SAN assembled and curated sequence data; TGV and JS performed analyses; TGV, JS, SAN and TS wrote the paper. The authors declare no competing interests.

## Abstract

We estimate the basic reproductive number and case counts for 15 distinct SARS-CoV-2 outbreaks, distributed across 10 countries and one cruise ship, based solely on phylodynamic analyses of genomic data. Our results indicate that, prior to significant public health interventions, the reproductive numbers for a majority (10) of these outbreaks are similar, with median posterior estimates ranging between 1.4 and 2.8. These estimates provide a view which is complementary to that provided by those based on traditional line listing data. The genomic-based view is arguably less susceptible to biases resulting from differences in testing protocols, testing intensity, and import of cases into the community of interest. In the analyses reported here, the genomic data primarily provides information regarding which samples belong to a particular outbreak. We observe that once these outbreaks are identified, the sampling dates carry the majority of the information regarding the reproductive number. Finally, we provide genome-based estimates of the cumulative case counts for each outbreak, which allow us to speculate on the amount of unreported infections within the populations housing each outbreak. These results indicate that for the majority (7) of the populations studied, the number of recorded cases is much bigger than the estimated cumulative case counts, suggesting the presence of unsequenced pathogen diversity in these populations.

**Significance Statement:** Since the beginning of the COVID-19 outbreak in late 2019, researchers around the globe have sought to estimate the rate at which the disease spread through populations prior to public health intervention, as quantified by the parameter *R*_0_. This is often estimated based on case count data and may be biased due to the presence of import cases. To overcome this, we estimate *R*_0_ by applying Bayesian phylodynamic methods to SARS-CoV-2 genomes which have been made available by laboratories worldwide. We provide *R*_0_ and absolute infection count estimates for 15 distinct outbreaks. These estimates contribute to our understanding of the baseline transmission dynamics of the disease, which will be critical in guiding future public health responses to the pandemic.

---

The novel coronavirus SARS-CoV-2 and the corresponding disease COVID-19 continue to spread at an alarming rate. As of the 27^th^ of August 2020, close to 9 months after its initial identification, over 24 million confirmed cases and nearly seven hundred thousand deaths have been reported globally. (1).

In order to understand the global threat this pandemic poses, it is necessary to accurately quantify the underlying transmission dynamics of the virus and, in particular, its basic reproductive number (2). This information is used to determine the likely future trajectories of individual outbreaks, and to retrospectively assess the impact of containment measures. Inference of the transmission dynamics is traditionally achieved using line list data (3) comprised of case confirmation times, locations and patient details, and this approach is being widely applied (4–7) by various groups around the world seeking to understand the current pandemic.

In particular, the *EpiForecasts* platform (8) is reporting frequently-updated results based on these methods as new line list data becomes available. While details vary between countries, these analyses indicate that the median estimates for the basic reproductive number for the populations studied in this report lie between 1.5 and 3. Our own monitoring of the reproductive number is updated daily using the latest confirmation, hospitalization, death, and excess death data with a focus on European countries (9) leading to similar results.

Despite the wide-spread application of such methods, the estimates produced by line list data alone are inherently susceptible to several biases and limitations (10–12). Firstly, the presence of pools of undiagnosed infected individuals, together with changes in testing methods and the extent to which testing is happening at all, can lead to misleading characterizations of the epidemic. Secondly, it is often impossible to discriminate between import cases and those attributable to local transmission based on line list data. This has the potential to produce overestimates of local transmission rates. Estimating rates and directions of transmission between geographic regions is similarly impeded. Thirdly, on their own, these data do not provide information about the state of outbreaks before the first recorded case.

Characterizing transmission dynamics is critical to the successful design of public health interventions. Thus, finding ways around potential biases and limitations when quantifying transmission dynamics is crucial. Fortunately, early testing efforts have been paralleled by significant efforts to sequence SARS-CoV-2 genomes from the initial outbreak and subsequent pandemic in “real time”. Many of the groups responsible for sequencing SARS-CoV-2 genomes have generously chosen to make them available immediately to the public research community via the GISAID platform (13). These data have been successfully used for the development of testing assays (14) and for learning about the molecular structure of the virus (15, 16). Importantly, the continued and widespread sequencing efforts has also enabled — in combination with phylodynamic methods (17, 18), independent, and potentially more robust, estimates of very early transmission dynamics.

Phylodynamic methods couple epidemiological models with models of sequence evolution, allowing us to estimate transmission dynamics based on the relationships between SARSCoV-2 genome sequences. Several studies have already made use of SARS-CoV-2 sequence data in a phylodynamic context. For example, Lai et al. (19) inferred early dynamics of the global effective reproductive number, using all available sequences at the date of publishing, obtaining an *R*_0_ estimate of 2.6, with a 95% credible interval [2.1, 5.1]. In contrast, Volz et al. (20) focused on a specific Wuhan-associated outbreak cluster and used a compartment model to also infer a basic reproductive number of 2.6, but with a 95% credible interval [1.5, 5]. Genomes have also been coupled with extremely detailed agent-based models to infer the probable sources of infection for specific COVID-19 cases within the Australian population (21).

In this paper we go further and infer the basic reproductive number (*R*_0_) for each of 15 distinct outbreaks distributed among 10 countries and the Diamond Princess cruise ship using phylodynamic methods. We use Bayesian model averaging to quantify the evidence for distinct *R*_0_ values as opposed to groups of outbreaks sharing *R*_0_ values. Finally, we provide Bayesian estimates of cumulative case counts over time for each of the outbreaks as ensembles of possible trajectories.

## Results

We used the *NextStrain* (22) platform to identify outbreak clusters for which sequence data exist, and selected only those sequences sampled prior to or just after the introduction of strong public health interventions in the associated locations (see Methods). (The Diamond Princess outbreak is an exception to this protocol, as the interventions were put in place immediately on the date corresponding to the first sequenced sample.)

We then applied a Bayesian phylodynamic framework (17), to co-infer *R*_0_ along with the probability of a infected person being included in our dataset, and the underlying phylogenetic trees for these clusters. This inference was done under the assumption of constant transmission rates (i.e. a constant rate birth-death process) for each cluster, with the sole exception of the Diamond Princess, where we allowed for the transmission rate to shift at the time of the onboard quarantine.

Figure 1 illustrates the posterior distributions for *R*_0_ associated with each of the outbreaks, together with the prior distribution for comparison. Interestingly, rather than a continuum of values, our analysis seems to isolate several distinct modes. The median posteriors for the majority of outbreaks lie between 1.4 and 2.9. However, the *R*_0_ values inferred for the two outbreaks associated with Iceland, the Welsh outbreak, a Washington State (USA) outbreak and the Diamond Princess outbreak have posterior median values ranging between 4 and 7. We used a Bayesian model averaging scheme to quantify the number of significantly distinct *R*_0_ values among all outbreaks, and found support for four distinct values (see figure S1 for the posterior distribution). The corresponding posterior distributions for the outbreak-specific *R*_0_ values generated by this model are shown in figure S2. A comparison of the preand post-quarantine effective reproductive number estimates for the Diamond Princess outbreak is shown in figure S3, and shows a significant drop in transmission rate following the im plementation of isolation measures. The proportion of infected individuals sampled for sequencing in each outbreak was also inferred as part of this analysis and these results are shown in figure S4.

**Fig. 1.**
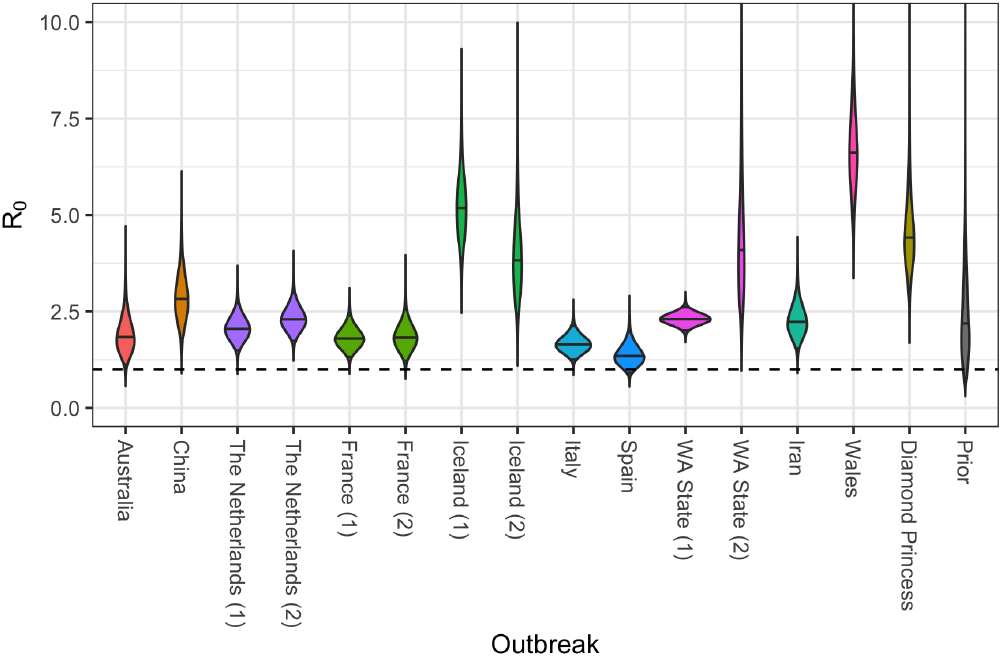
Posterior distributions for reproductive numbers for outbreaks considered in this study. Solid horizontal lines represent median values; the dashed horizontal line represents the threshold between exponential growth and decline of outbreak.

Birth-death phylodynamic results are dependent not only on the genomic data, but also on the distribution of sample collection dates. In fact, we find that in this instance, the sample collection dates carry *most* of the information regarding *R*_0_. We demonstrated this by running an additional set of phylodynamic analyses in which the genomic sequences were treated as unknown. Additionally, we applied both a simplistic linear regression approach (see Methods) and an established traditional approach (12) to the cumulative sequence counts. The results of these alternative analyses are summarized in figure S5 and—in many cases—show relatively close agreement, albeit with slightly less certainty in the estimates than those shown in figure 1.

Given this dominating effect of the sampling times, it is natural to consider how sensitive our results are to the assumption that the sampling rate and reproductive number are fixed over the time period of each outbreak. We thus performed a separate set of analyses in which these quantities were allowed to change at a point at the center of the sampling window of each outbreak (excluding the Diamond Princess outbreak). The resulting *R*_0_ estimates, presented in figure S8, show no major change in the results compared with those in figure 1, with the exception of the Netherland (1) and WA States (1) outbreaks which suggest higher *R*_0_ values. In order to investigate how much our results are impacted by the prior, we repeated the fixed-rate analyses with a broader prior on *R*_0_. This broad prior did not qualitatively change the results compared to our main analysis (figure S9).

Inferred cumulative case count trajectories for the Washington State and Diamond Princess outbreaks are shown in figure 2 alongside the daily number of confirmed cases as reported by the Center for Systems Science and Engineering at Johns Hopkins University (23), to which we have applied a 10 day offset in order to account for the estimated delay between infection and case confirmation (9). In the cases where two outbreaks are associated with the same location, the inferred case counts are combined. Similar case count trajectories for the remaining populations are provided in figure S6. The posterior distributions for case counts at the time of the most recent genome sample are shown for all populations in figure 3.

**Fig. 2.**
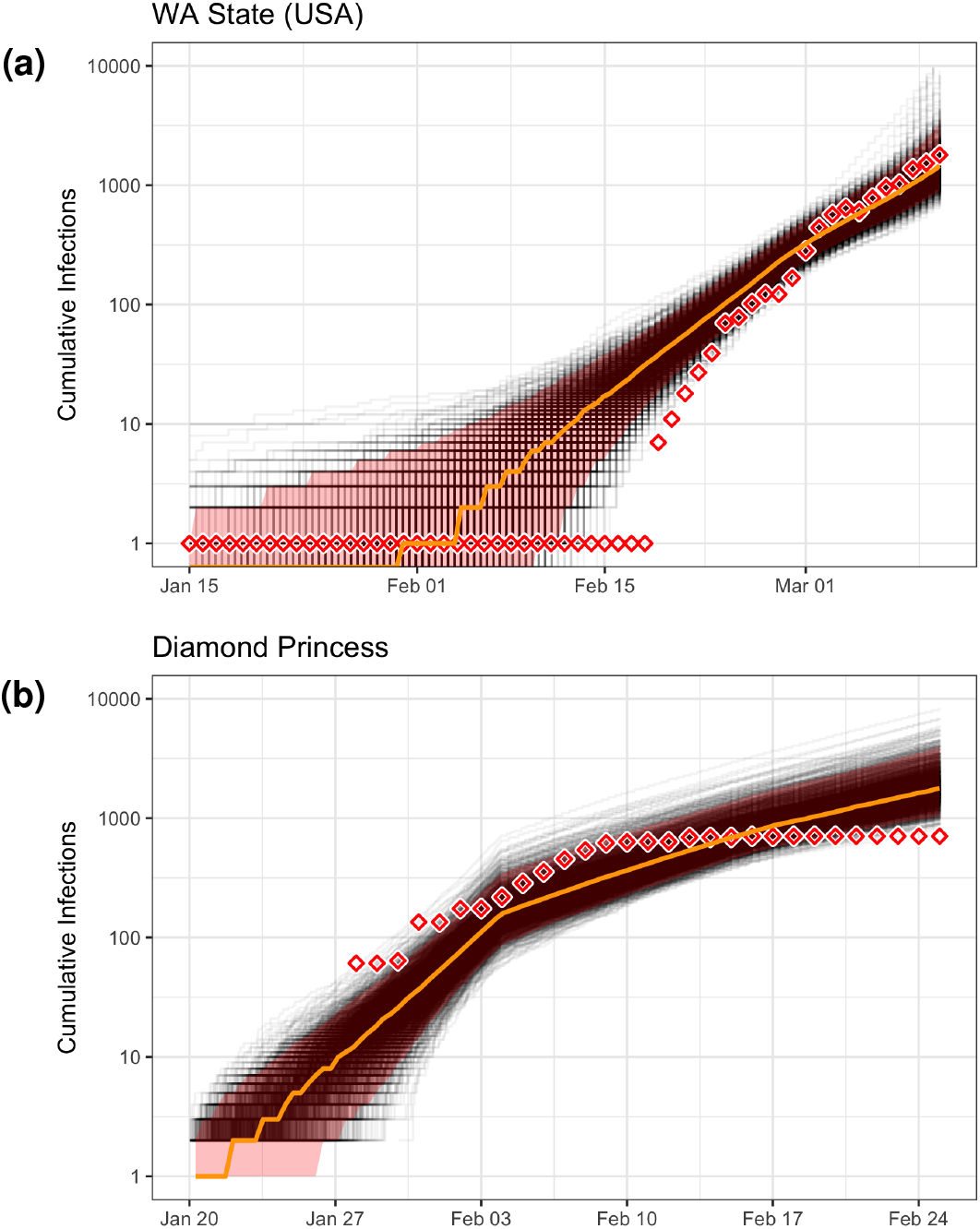
Inferred cumulative case count trajectories for (a) Washington State, USA, and (b) the Diamond Princess cruise ship. They are shown together with the corresponding recorded case counts (diamonds) in each population as recorded by Dong, Du and Gardner (23), which are offset by 10 days to account for the delay between infection and case confirmation (9). Note that these inferences concern only those cases associated with the specific outbreak from which the sequence data are drawn, as detailed in the discussion section. The true cumulative case counts may have been much higher. (Inference for remaining outbreaks are shown in figure S6.)

**Fig. 3.**
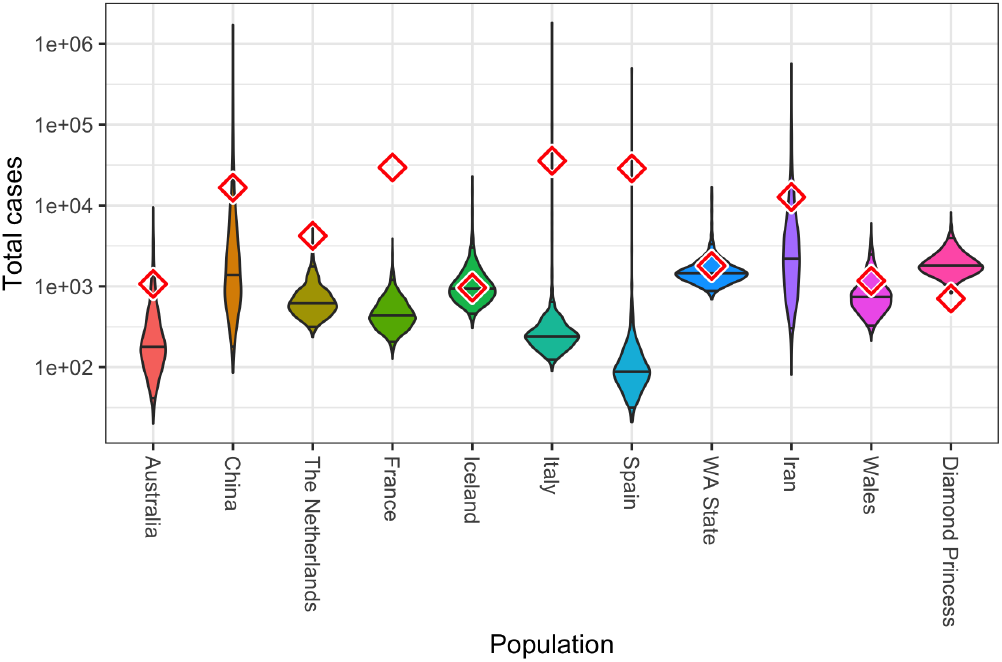
Estimates of cumulative case counts obtained from phylodynamic analyses, with diamonds indicating recorded counts obtained from Dong, Du and Gardner (23), offset by 10 days to account for the delay between infection and case confirmation (9). The counts are for the date of the final genome sample considered in each population. We note that we have likely analyzed only a subset of the total number of outbreaks which were circulating in each country.

## Discussion

Our central result is that prior to strong public health interventions, the majority (10) of the outbreaks studied seem to have grown at rates with median *R*_0_ values ranging between 1.4 (Spain) and 2.8 (China).

The specific case of the Diamond Princess is interesting, as the details of this outbreak are well known and, at least for the time period affecting our analysis, the population involved was strictly isolated (i.e. we can say with a high degree of certainty that no immigration or emigration occurred). In this case, we believe the high pre-intervention *R*_0_ estimate reflects a real elevated infection rate caused by unchecked transmission within the relatively confined on-board environment.

The remaining outbreaks to which higher *R*_0_ values are attributed are limited to those with the shortest sampling windows (see figure S7). Given the strong role played by sample times in these inferences, it is therefore possible that these values are the result of bias due to sampling model misspecification, and that this problem is exacerbated by the short sampling windows involved. The sampling model used for these outbreaks assumes that samples accumulate at a rate proportional to the number of infectious cases for the duration over which samples are available. We showed that allowing for a single shift in sampling rate and *R*_0_ during the outbreak did not result in much lower *R*_0_ values for these remaining outbreaks.

Another potential source of upward bias on *R*_0_ is the process of outbreak selection. We necessarily restrict our attention to outbreaks for which sufficient data exist to provide statistical signal. This restriction may have the effect of selecting for steeper outbreak trajectories. Since the birth-death models under which we perform the inference do not account for this conditioning, these steeper trajectories will be interpreted as evidence for larger *R*_0_, even when the increased gradient is simply the result of demographic noise in the growth of the epidemic. Including appropriate conditioning in phylodynamic inference to guard against this kind of bias will be the focus of future research.

Given that most of information content in the genome sequence data analyzed seems to come from sampling times, it is natural to wonder whether the phylodynamic approach offers additional insights on these outbreaks. The obvious answer to this challenge is that, genomic data allowed us to identify outbreaks in the absence of contact tracing data, which is often not available for study. Furthermore, even though the impact of the phylogeny *within* each identified outbreak on the inferred epidemic parameters was negligible, the application of phylodynamic methods yield information about the total number of cases through time, including those which have gone undetected.

We emphasize, however, that extreme care must be taken when interpreting both these inferred and clinically confirmed case counts as representative of the true underlying case load. Firstly, our inferences correspond to the number of cases associated only with the specific outbreaks from which the genomic data originate. It is entirely possible that additional outbreaks, from which we do not have genetic data, occurred within a given population during the time periods considered. These cryptic outbreaks could contribute to the confirmed case counts but would be absent from our phylodynamic inference. Secondly, the confirmed case numbers themselves can only provide a lower bound on the true number of cases in a population. Taken together, these points imply that the larger of the phylodynamically-inferred case counts and the corresponding confirmed case counts provide a lower bound on the true number of cases within each population.

We highlight in this paper the importance of SARS-CoV-2 genomes for quantifying transmission dynamics. In particular, we provide estimates for the basic reproductive number which are complementary to classic epidemiological studies. Our phylodynamic analyses of SARS-CoV-2 genomes confirm the *R*_0_ estimates for Wuhan (5) and provide estimates for 15 outbreaks around the world for which classic epidemiological methods are problematic due to the difficulty of disentangling introductions from local transmissions. Going forward, we are convinced that SARS-CoV-2 genomes will become useful for quantifying changes in transmission rates (for instance, using the BDSKY model of Stadler et al. (17)) and become essential for evaluating the importance of local transmission versus imports (for instance, using the multi-type birth-death model of Kühnert et al. (24)). The latter is in particular important after the end of lock-down measures which we currently experience in many European countries. Indeed, for patients whose infection is not traceable, it is the genomes which contain valuable information for linking them into the transmission chain and thus quantify transmission dynamics.

## Materials and Methods

### Outbreak identification and sample selection

The birth-death models we employ assume that genome samples are taken uniformly at random from the infectious population during the early, exponential growth phase of each outbreak. Since our analysis is necessarily retrospective rather than prospective, we devised two strategies to approximate such a sampling scheme using publicly-available samples from GISAID (13). For sparsely-sampled, un-sampled, or clearly non-uniformly sampled outbreaks (Italy, Iran, and China before the quarantine of Wuhan, respectively), we included sequences from cases that were exposed in the region of interest and subsequently traveled abroad, where they were then diagnosed and sampled. (The sequences attributed to the Iranian outbreak, for example, are all travel cases isolated and sequenced in Australia (25).) For more densely-sampled outbreaks (France, Iceland, the Netherlands, Spain, Wales, and Washington State, USA), we analyzed samples that were exposed and sampled within the region of interest. For these outbreaks, we considered only samples that clustered together with other samples from the same region in a phylogenetic tree of the global pandemic (22). This was done in order to sample primarily within-region transmission events.

#### Sample acquisition and curation

We downloaded all sequences available on GISAID (13) on April 1^st^, 2020. After quality-filtering this sequence set, we aligned the sequences, built a phylogenetic tree, and identified regional outbreak clusters within the tree. Sequence quality-control, alignment, and tree building were all performed using the Nextstrain pipeline adapted to SARS-CoV-2 (26).

We first filtered the available sequences to exclude sequences shorter than 25,000 base pairs, sequences with imprecise sampling dates, known re-samples of the same case, low-quality sequences (as determined by Nextstrain), and all but one sequence from known epidemiologically-linked cases. We note that our knowledge of which samples come from epidemiologically-linked cases (as identified by Nextstrain and gleaned from media reports) is far from exhaustive. Whenever we were able to access this information we used it to exclude non-randomly sampled sequences, but in many cases the relevant information was either not collected or not readily accessible.

#### Alignment and outbreak detection

After these filtering steps, we aligned the remaining sequences to a reference genome generated from an early COVID-19 patient in Wuhan (GenBank accession number MN908947) (27). SNPs in the first 130 sites, last 50 sites, and at sites 18529, 29849, 29851, and 29853 were masked from the alignment because they are likely sequencing artifacts (26).

We built a maximum-likelihood phylogenetic tree using this alignment. We then picked clades from this tree where sufficient (≥9) samples from the same region clustered together. We assume that these clusters represent primarily within-country transmission events rather than introductions from abroad.

Exceptionally for the Itay, Iran, and China outbreaks we additionally identified samples from cases that were presumably exposed to the virus in these regions but were sampled abroad (travel cases). The data set for Italy included sequences from both non-travel and travel cases, while those for China and Iran were composed exclusively of sequences from travel cases. This exposure information comes from metadata available on GISAID and Nexstrain, as well as information provided by sequencing centers and in media accounts.

#### Sample set truncation

To limit sampling to the early, exponential growth phase of each regional outbreak, we truncated sampling based on the dates of major public health interventions (Table S1). We retained only samples collected before or on the date of theses public health interventions, with the exception of the Iran, Iceland, and Spain outbreaks. For these outbreaks, we extended the time cutoff so that the sample size was not prohibitively small. (The extension for Iran was 11 days, for Iceland it was 2 days, and the cutoff for Spain was extended by 1 day, as shown in Table S1.) Since the transmission events leading to sampled cases likely happened at least a few days before sampling, these cutoffs should, for the most part, be conservative.

### Phylodynamic analyses

#### Main analyses

Sequence alignments were analyzed jointly as part of a single Bayesian phylodynamic analysis using the BDSKY package (17) of BEAST 2 (28), using a single HKY+Γ substitution model with a strict clock rate fixed to 8 10^4^ substitutions/site/year (following Nextstrain (22)). The tree *T*^(*c*)^ corresponding to each outbreak cluster *c* was assumed to be produced by a birth-death process with reproductive number 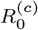, sampling proportion *s*^(*c*)^ and become uninfectious rate *δ*. In each case, the sampling proportion for the outbreak was assumed to be zero before the first included sample for that outbreak. In the special case of the Diamond Princess outbreak, a second (effective) *R*_0_ value was associated with the days following the on-board intervention. All *R*_0_ values were assumed to be independent and given a LogNormal(0.8, 0.5) prior. The time between the start of the birth-death process associated with each outbreak and the time of the most recent sample for the same outbreak was given a LogNormal(−2, 0.8) prior. The value of the become uninfectious rate *δ* was fixed to 36.5, equivalent to an expected time until becoming uninfectious for each individual of 10 days. (This is in line with the estimates of the latent and infectious periods provided by Li et al. (4), and follows the assumptions used by Sciré et al. (9).)

A second analysis was run with an identical model configuration to the first analysis, aside from its use of Bayesian model averaging to quantify the number of distinct 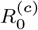 values needed to describe the outbreaks. This was done by applying a Dirichlet process prior (DPP) to the vector 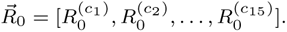 Following the prescription of Dorazio (29), a Gamma hyperprior was applied to the intensity parameter of the DPP such that the implied prior distribution for the number of unique elements of 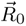 was as close to uniform as possible. The base distribution of the DPP was chosen to be LogNormal(0.8, 0.5). The prior for each the sampling proportion was chosen to be Beta(1, 4), which prioritizes low sampling probabilities without completely excluding higher probabilities.

#### Sensitivity analyses

We ran two additional analyses to determine the sensitivity of our conclusions to the model assumptions. Firstly, to test the robustness with respect to changes in the *R*_0_ priors, we ran a separate analysis using a Unif(0,10) prior for each 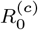 parameter. Secondly, we ran an analysis in which both 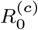 and *s*^(*c*)^ were allowed to change once during each outbreak, at a time midway between the first and last sample assigned to that outbreak.

#### Sample-date only analyses

In order to assess the relative impact of the sequence data on these 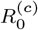 estimates, another joint phylodynamic analysis was performed using the same setup as the first, but without any sequence data.

Additionally, a simple regression inference of the 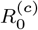 was conducted by assuming that the number of active infections associated with each outbreak grew according to the deterministic function 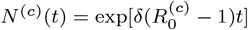. This implies that the logarithm of the cumulative number of samples grows linearly at the rate 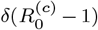, which we then fit to the empirical cumulative sample numbers from each outbreak.

In order to test the robustness of the phylodynamic estimates of the outbreak-specific 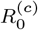 values, we applied EpiEstim (12) to the same sample time distributions used for the regression analysis. In these analyses, 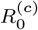 was assumed to be constant through time in each outbreak. A serial interval of mean 4.8 days and standard deviation 2.3 days was used (30).

#### Case count trajectory inference

Inference of cumulative case count trajectories was achieved by applying the particle filter algorithm implemented in EpiInf (31) to the outbreak-specific tree and parameter posteriors produced by the corresponding BDSKY analyses.

## Data Availability

All of the files necessary to replicate the analyses presented in this manuscript are made available, as detailed in the manuscript, including accession numbers for genomic data from the GISAID sequence database.

https://github.com/tgvaughan/R0-manuscript-materials/

## Data availability

The sequences used in this study are distributed via GISAID (https://gisaid.org). The acknowledgements table available at https://github.com/tgvaughan/R0-manuscript-materials/blob/aster/sequences/GISAID_Acknowledgement_Table.csv lists the accession numbers for the sequences associated with each cluster, together with the names of the institutions and authors who generously contributed the sequences.

The BEAST 2 XML files used to perform the phylodynamic analyses, together with the R scripts used for post-processing, are available from https://github.com/tgvaughan/R0-manuscript-materials/.

## ACKNOWLEDGMENTS

We thank the numerous institutions and authors who generously made SARS-CoV-2 genomes available for public research via the GISAID platform. TGV, JS, SAN and TS thank ETH Zürich for funding.

